# Is there a role for RDTs as we live with COVID? An assessment of different strategies

**DOI:** 10.1101/2022.09.30.22280569

**Authors:** Gabrielle Bonnet, Anna Vassall, Mark Jit

## Abstract

**Introduction:** By 2022, high levels of past COVID-19 infections, combined with substantial levels of vaccination and the development of Omicron have shifted country strategies toward burden reduction policies. SARS-CoV-2 rapid antigen tests (RDTs) could contribute to these policies by helping rapidly detect, isolate and/or treat infections in different settings. However, the evidence to inform RDT policy choices in LMICs is limited.

**Method:** We provide an overview of the potential impact of several RDT use cases (surveillance; testing, tracing and isolation without and with surveillance; hospital-based screening to reduce nosocomial COVID; and testing to enable earlier/expanded treatment) for a range of country settings. We use conceptual models and literature review to identify which use cases are likely to bring benefits and how these may change with outbreak characteristics. Impacts are measured through multiple outcomes related to gaining time, reducing the burden on the health system, and reducing deaths.

**Results:** In an optimal scenario in terms of resources and capacity and with baseline parameters, we find marginal time gains of at least a week through surveillance and testing tracing and isolation with surveillance, a reduction in peak ICU or ICU admissions by 6% or more (hospital-based screening; testing, tracing and isolation), and reductions in COVID deaths by over 6% (hospital-based screening; test and treat). Time gains may be used to strengthen ICU capacity and/or boost vulnerable individuals, though only a small minority of at-risk individuals could be reached in the time available. The impact of RDTs declines with lower country resources and capacity, more transmissible or immune-escaping variants and reduced test sensitivity.

**Conclusion:** RDTs alone are unlikely to dramatically reduce the burden of COVID-19 in LMICs, though they may have an important role alongside other interventions such as vaccination, therapeutic drugs, improved healthcare capacity and non-pharmaceutical measures.

**KEY MESSAGES:**

- **What is already known:** Important shifts in the way COVID is addressed, from efforts to reduce COVID transmission toward burden reduction, have led to question the role of testing, in particular SARS-CoV-2 rapid antigen tests (RDTs), both in countries that used them extensively and in those in which RDTs were never scaled-up, but the evidence to inform RDT policy choices in LMICs is particularly limited.
- **What this study adds:** This study provides an overview of multiple RDT use cases and their potential impacts to gain time, reduce health system burdens and reduce deaths. It shows the contrast between high- and low-resource and capacity settings, and how some use cases (surveillance, hospital-based screening, and testing associated with early/expanded treatment) may retain higher benefits, at least with regard to early warning and hospital burden, than the use of RDTs for testing, tracing and isolation.
- **How this study might affect research, practice or policy**: The study highlights that RDTs alone are unlikely to dramatically reduce the burden of COVID-19 in LMICs, and that their role may be best understood as complementary to other interventions.

## INTRODUCTION

As of mid-2022, the COVID-19 pandemic had caused over half a billion reported cases and 6 million reported deaths worldwide [1], with the actual death toll expected to be three times greater [2]. The high level of past infections, particularly associated with the development of the Omicron variant, combined with mass vaccination (60% of the world population vaccinated by mid-2022 [1]) and the development of new therapeutics [3], have led to a shift in policies, from efforts to reduce COVID-19 transmission toward efforts to reduce its burden. Since COVID-19 has become endemic and new variants that escape natural immunity are likely to continue to emerge, any burden reduction policies may need to be sustainable indefinitely.

SARS-CoV-2 rapid antigen tests, also known as rapid diagnostic tests (RDTs), provide results within 20-30 minutes, do not require laboratory equipment and are relatively inexpensive [4]. Up to 2022, their use, primarily for self-testing, has often been promoted in high-income countries to people at risk of SARS-CoV-2 infection to identify candidates for isolation and hence reduce transmission. They have been less widely used in most low-and-middle-income countries (LMICs) though there has been support for scaled-up, equitable RDT access [5 6]. With global focus increasingly shifting to burden reduction, countries are rethinking RDT policies. For example, the United Kingdom has gone from heavy reliance on RDTs to stopping free RDT provision [7]. The evidence to inform LMICs’ policy choices in this changing context remains limited. Models highlight that higher transmission levels reduce RDT effectiveness while combining RDT use cases with interventions that reduce transmission increases effectiveness [8]. Some models further [8-10] suggest that large-scale community screening or saturating testing demand through RDTs may be cost-effective provided isolation and sensitivity levels are sufficient. This may however be impossible to achieve in many low-resource settings.

The purpose of this paper is to provide an overview of the potential impact of various RDT use cases for a range of country settings, integrating data from literature reviews in broad conceptual models with supporting analytical components, to identify which use cases are likely to bring benefits and how these may change with outbreak characteristics. This should help countries focus on the strategies that may require further context specific modelling of impact and value for money before deployment. We focus on a number of use cases identified in WHO [11] and Africa CDC [12]’s guidance documents.

## METHODS

### Overview

The use cases explored in this paper: 1) RDT-supported surveillance; 2a) testing, tracing and isolation (TTI); 2b) TTI combined with surveillance; 3) hospital-based testing to control nosocomial COVID; and 4) RDTs to prompt improved/earlier treatment of those that may benefit from it, are shown in Figure 1, alongside their impacts during a COVID outbreak. These are measured through six different outcomes related to gaining time, reducing the burden on the health system, and reducing deaths.

**Figure 1:**
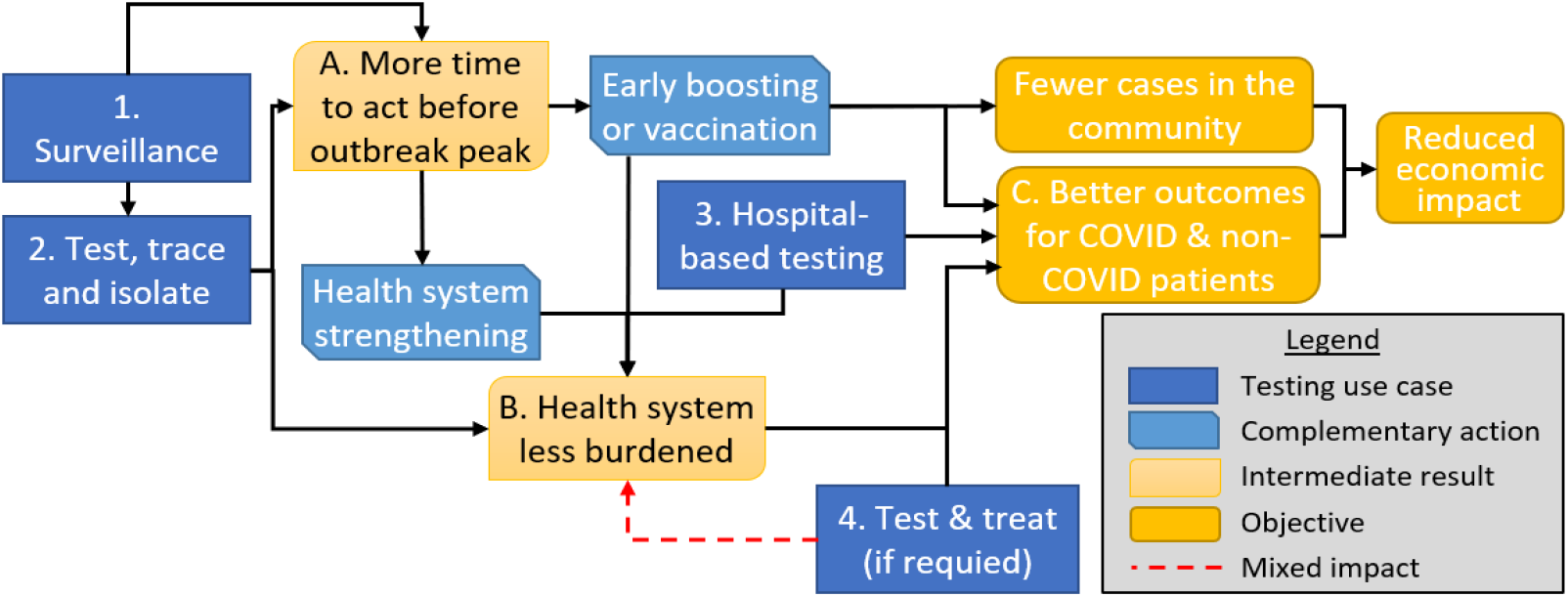
Potential roles of RDTs within burden reduction strategies.

### Model assumptions

#### Outcome metrics

We use the following outcomes to evaluate the success of different use cases:

A1) Time available for vaccination boosting (relevant to use cases 1, 2a and 2b). This is defined as the time between outbreak detection (see use case 1 below) and one week (since it takes around a week [13 14] for boosting protection to kick in) before the share of persons susceptible to COVID is halved as compared to the beginning of the outbreak. The outbreak starts when infections are first introduced in an otherwise uninfected community – at that point a proportion *s*_*0*_ of the population is susceptible to the disease.

We further translate time gained for boosting into the share of 60+ and 80+ years old (common priority groups for vaccination) who could be boosted in that time [1] assuming two vaccination speeds: the initial speed at which the population was vaccinated, and the speed attained once 1% of the population had been reached.

A2) Time available for ICU capacity increases (relevant to use cases 1, 2a and 2b). It is defined as the time between outbreak detection and the moment half of the ICU bed-days that would be used in an unmitigated outbreak have already been used.

B1) Percentage reduction in the height of the peak in ICU demand (relevant to use cases 2a and 2b). This indicator is further translated into a reduction in what we term ‘unmet needs’ (based on a given ICU capacity), or the total of ICU bed-days demand exceeding a certain threshold termed ‘ICU capacity’.

B2) Percentage reduction in ICU admissions (relevant to use case 3). B3) Percentage reduction in hospital admissions (relevant to use case 4).

C) Percentage reduction in country-wide COVID-related deaths (relevant to use cases 3 and 4).

#### Outbreak and test characteristics

In our base scenarios, the COVID outbreak has R_0_=10 (approximate value for the Omicron variant [15]), an initial share of immune individuals, from vaccination or past infection, *s*_*0*_ = 50%, and average latent and infective periods of 4 and 6 days respectively [16]. Here *s*_*0*_ could consist of a mixture of full immunity for those infected with the same variant and partial immunity for those vaccinated against or infected with a mismatched variant. We further assume RDT sensitivity is 80% (minimum acceptable in WHO guidelines [11]). RDT results are assumed to be immediate. For use cases for which we rely on data or models developed by other researchers, we choose the figures closest to our base assumptions. We address alternative outbreak and test parameters in the sensitivity analysis.

##### Scenario design

The impact of each RDT use case is assessed for multiple scenarios, summarized in Table 3 into six country “archetypes” representing country situations at different resource and capacity levels. These are associated with corresponding levels of RDT scale-up and values for other intervention and contextual parameters such as isolation, tracing, treatment availability and case/death identification. We assume in the lowest-resource scenario that 5% of symptomatic cases with RDTs are reached (more than a 10-fold increase over 2021 case ascertainment levels in low-income countries as per [2]), that in the following scenario (patterned on lower-middle-income countries) 10% are reached, then 20-40% (for scenarios patterned on upper-middle income countries) then 60-80% (for scenarios patterned on high-resource settings). Each of these scenarios is additionally associated with a set of use-case specific assumptions that are detailed in Appendix A.

### RDT use cases

#### Use case 1: Surveillance

In this use case, we assume that testing of individuals with COVID-19-like symptoms enabled by RDT use will allow for early detection of a COVID-19 outbreak (through an increase in confirmed cases) so that vaccination and/or ICU capacity increases can be initiated earlier (outcomes A1 and A2). Outcomes with early detection are compared to outcomes with no or poor surveillance, assuming that without surveillance outbreaks are only detected when COVID-19-related hospital admissions increase.

We use as an example the South Africa surveillance system; this may be an attainable model for some LMICs. We use four and two weeks as the time between early (respectively late) detection and the peak of the outbreak. These represent a rounding of the South Africa 2021 Omicron outbreak timeline: 26 days elapsed between the trough in reported cases [17] and the estimated peak in infections [2] and 15 days between reports of increasing hospital admissions [18] and peak infections. We then derive time available for boosting and for ICU capacity building using a simulation model, which we also apply in use cases 2a and 2b. This model starts at t = 0 when a new COVID-19 variant is introduced in an otherwise uninfected population, with one infected individual per million inhabitants. The model has a well-mixed population representing susceptible, exposed, infected and hospitalized (general ward and ICU) cases. The initial share of susceptible individuals is *s*_*0*_ = 50%. Details of the model structure and approach are presented in Appendix A. We then use data on past vaccination [17] to estimate the share of 60+ and 80+ years old that can be boosted in the time available.

#### Use cases 2a and 2b: TTI

There are two use cases involving TTI: 2a and 2b. In both use cases, we assume that testing of symptomatic cases and their contacts allows isolation of positives and (at least in some scenarios) tracing of their contacts, reducing transmission. In turn, transmission reduction increases time available for both boosting and the health care response and also reduces peak ICU demand (outcomes A1, A2 and B).

In use case 2a, we compare TTI with late outbreak detection (no surveillance) to late outbreak detection alone. In use case 2b, we compare TTI with early outbreak detection (surveillance, as per use case 1) to early outbreak detection alone.

In both 2a and 2b, we assume that α = 35% of cases are asymptomatic [19], with an infectious period *T*_*asym*_ = 7 days and relative infectivity (the reduction of the effective contact rate for asymptomatic cases as compared to symptomatic ones) *f* = 0.5, while preclinical and clinical infectiousness for symptomatic cases last *T*_*pre*_ = 2.4 and *T*_*clin*_ = 3.2 days, respectively [16]. Delay between first symptoms and testing is set at *T*_*test*_ = 0.5 days, reflecting time to decide to test and/or procure a kit. Transmission reduction following isolation is φ = 70%, the share of transmission outside the home [20] (isolation from household members is difficult, particularly in poor settings). The computation of the reduction in transmission following testing, tracing and isolation is detailed in Appendix B.

Finally, we use the model described in use case 1 and Appendix A to translate transmission reduction into time gained for boosting, time gained to strengthen ICU capacities, a percentage reduction in peak ICU demand and a reduction in ICU ‘unmet needs’ (given capacity) for different levels of testing, tracing and isolation without (use case 2a) and with (use case 2b) surveillance. As in use case 1, we further use data on past vaccination [17] to estimate the share of 60+ and 80+ years old that can be boosted in the time available.

#### Use case 3: Testing in health facilities

In this use case, we assume that screening of staff and/or hospitalized individuals with RDTs, in combination with routine PCR use, can reduce nosocomial COVID prevalence as compared to routine PCR use alone. Nosocomial COVID itself creates heightened risks for vulnerable hospitalized individuals and puts a strain on hospital capacity through patient and staff illness. Reducing nosocomial COVID reduces deaths in vulnerable individuals (outcome C) and the burden on the health system through a reduction in the number of people admitted to ICU (outcome B2).

For this use case, we assume that nosocomial COVID prevalence is 20% before intervention (exploring a range of 8-46%), that the relative risk of ICU admission for nosocomial cases vs. hospitalized community-acquired COVID is 0.74 (0.50-1.08) while the relative risk of death is 1.3 (1.005-1.683) and that the marginal reduction in incident cases through RDT screening plus PCR as compared to PCR alone is 35.3-49.2%. Nosocomial COVID prevalence is based on a review undertaken early in the pandemic (considering only studies with a sample >100) [21] and recent information about prevalence with the Omicron variant [22-24]. Risks of ICU admission and death reflect a 2021 systematic review [25], which results were supported by later studies [26-29]. Finally, the impact of healthcare-facility screening is based on simulations in [30] for the high community incidence scenario. These results use lower RDT sensitivity than in our base assumption, require two rounds of patient and worker testing in two weeks in reaction to an outbreak and the capacity to isolate infected cases.

#### Use case 4: Testing and treatment

In this use case, we assume that increased RDT access leads to changes in care opportunities for high-risk individuals with mild or moderate disease that would be otherwise only be treated if and when they develop more severe symptoms, allowing them to access early treatment (e.g., with antivirals, monoclonal antibodies or facility admission). In turn, early treatment of high-risk cases leads to a reduction in the risk of hospital admission (outcome B3) and death (outcome C).

We assume that, as they are scaled up, RDTs progressively benefit firstly the most advantaged individuals (who would have access to rapid PCR testing and optimal care even in the absence of RDTs), and only after that people who would access care only if they developed severe symptoms. The last to be reached would be those without any access to care, even in the case of severe/critical disease.

Finally, we assume that, without improved care, the risk of hospitalization for high-risk patients is between 1.5 and 6% while with it, it is 0.9%. We also assume that mortality risk is reduced by 33-100% and that 25% of hospitalized patients and 50% of hospital deaths are high-risk [3 31-33]. The details of the formula and sources used for this use case and its justification are in Appendix C.

#### Sensitivity analysis

In the sensitivity analysis, we consider different outbreak characteristics, exploring scenarios with a different effective contact rate, initial share of susceptible individuals, and infectious and latent periods. We also discuss the impact of halving test sensitivity, in line with the impact of the Omicron variant on the sensitivity of several RDTs [34].

To assess the consequences of changes in variant characteristics on use case 1, we assume for simplicity that the outbreak is detected when infections exceed a fixed level *i*_*d*_ (for early detection enabled by surveillance), or when hospital admissions exceed a certain level *h*_*d*_ (for late detection in the absence of surveillance). *I*_*d*_ and *h*_*d*_ are set so that, for baseline outbreak characteristics, the delay between outbreak detection and peak infection rates be two and four weeks when infections vs. admissions are used respectively, in accordance with the assumptions for that use case. This corresponds to *i*_*d*_ = 0.087% and *h*_*d*_ = 0.042%. Meanwhile, if test sensitivity is halved, the infection level that can be detected with surveillance (assuming unchanged test specificity) doubles. The impact of changes in outbreak characteristics and RDT sensitivity is then simulated using the model in Appendix A while some mathematical formulas are provided in Appendix D.

For use cases 2a and 2b, we combine two effects: 1) the impact of a change in variant characteristics or test sensitivity on the time gained for boosting, time gained for ICU capacity building, and reduction in peak ICU demand, for a given level of advance notice 2) the impact of a change in the level of advance notice brought about by a change in outbreak characteristics or test sensitivity, and described in relation with use case 1. These are estimated again using both the simulation model (Appendix A) and, for some of these values, mathematical formulas (appendix D).

For use case 3, since we rely on the results of [30], we discuss the implications of the sensitivity analysis developed in that paper. We finally discuss the implications of variant and test sensitivity changes on use case 4 (these are more straightforward than for other use cases).

## RESULTS

### Use case 1: Surveillance and time available for boosting or ICU capacity building

Using the model in Appendix A, we estimate that 50% of the people susceptible at the start of the epidemic have already been infected 3.7 days before infections peak. In the no surveillance (resp. surveillance) scenario, the outbreak is detected around 2 (resp. 4) weeks before infections peak (see methods). Since time available for boosting is defined as the time between outbreak detection, on the one hand, and one week before the share of persons susceptible to COVID in the outbreak is *s*_*0*_/2, on the other, in the no surveillance scenario, time available for boosting is 3.3 days (14-3.7-7) and with surveillance, it is 17.3 days. We translate this time into a share of 60+ and 80+ years old that can be boosted in different scenarios (see Appendix B). Should 60+ (resp. 80+ years old) be targeted at the initial vaccination speed, the median share that can be boosted in LMICs with surveillance is 3.6% (resp. 35.5%) vs. 0.4% (resp. 4.6%) without. Should countries be able to boost at the speed they had reached after vaccinating 1% of their population, these figures would be 12.5% (resp. over 100%) with surveillance vs. 2.4% (resp. 26.3%) without.

We then focus on time available for ICU capacity increase. There are 19.2 days between peak infection and the moment half of all ICU bed-days required during the outbreak have already been used. Hence, time available for ICU capacity strengthening without surveillance would be 33.2 days, with surveillance it is 47.2 days.

### Use case 2: Testing, tracing and isolation (with or without surveillance)

Transmission reduction associated with different TTI scenarios was computed using the formulas in the methods and Appendix B. Values range from less than 1% reduction in low scenarios with 5-10% tested through RDTs and 25% isolating, to 24% for 80% testing, 75% isolating and 75% traced. Appendix B provides transmission reduction for a large range of scenarios.

#### Outcome A: Time gained

Table 1 provides the marginal increase in time available for boosting resulting from the use of TTI vs. no TTI. This increase depends on whether surveillance is present (use case 2b) vs. not (use case 2b). The marginal increase in time available for ICU capacity strengthening is very similar (Appendix B).

**Table 1:**
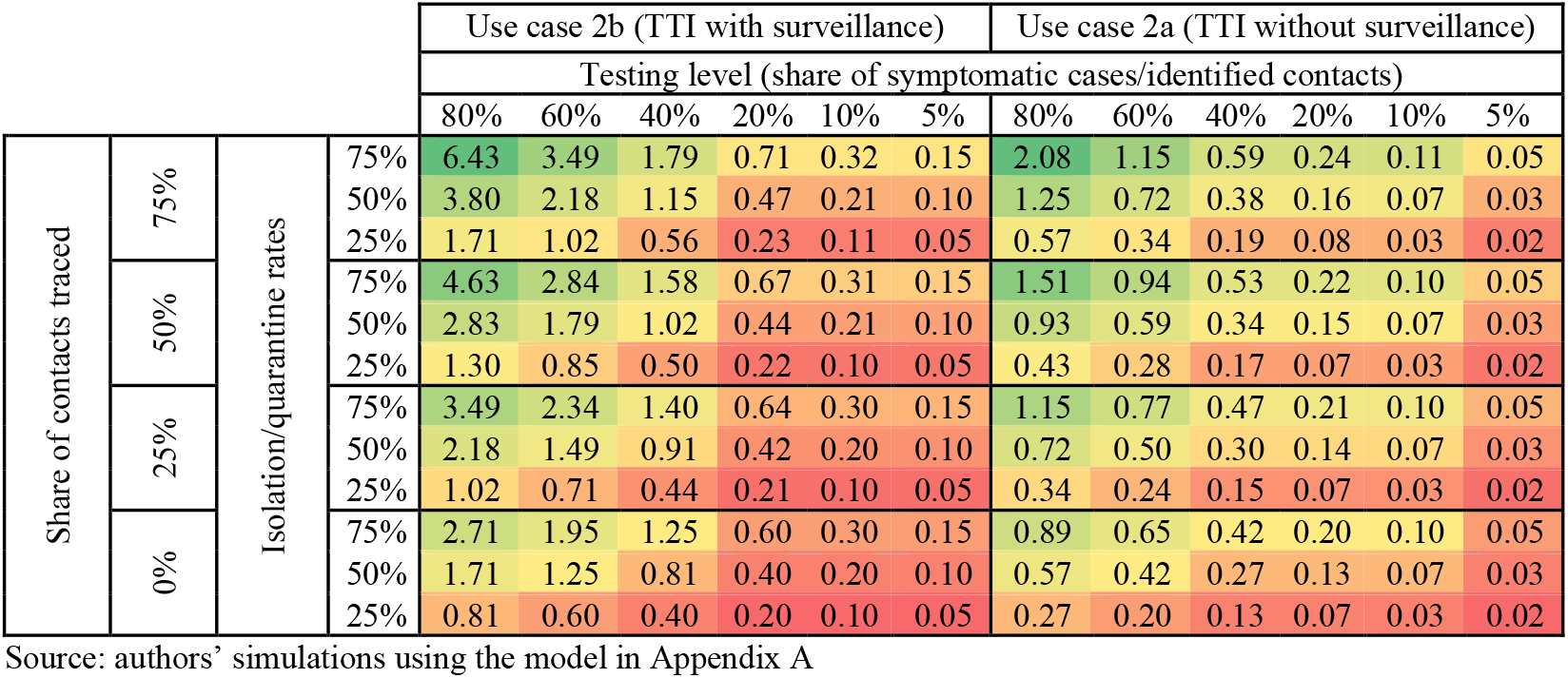
Time gained for boosting (in days) through TTI with different levels of advance warning

Adding this time to time available for action through surveillance alone (Use case 1), we compute the share of 60+ and 80+ years old that can be boosted in different scenarios (Appendix B). Should 60+ (resp. 80+ years old) be targeted at the initial vaccination speed, the median share that can be boosted in LMICs in the best scenario (optimal TTI + surveillance) is 6% (resp. 58%). Should countries be able to boost at the speed they had reached after vaccinating 1% of their population, these figures would be 17% (resp. over 100%).

#### Outcome B: Reduction in peak ICU needs

Table 2 translates transmission reduction into a percentage reduction in peak ICU needs. Note that transmission reduction does not so much reduce the total number of ICU cases as it changes the shape of the outbreak, hence reducing peak needs, which reduces the risk of the health system being overwhelmed.

**Table 2:**
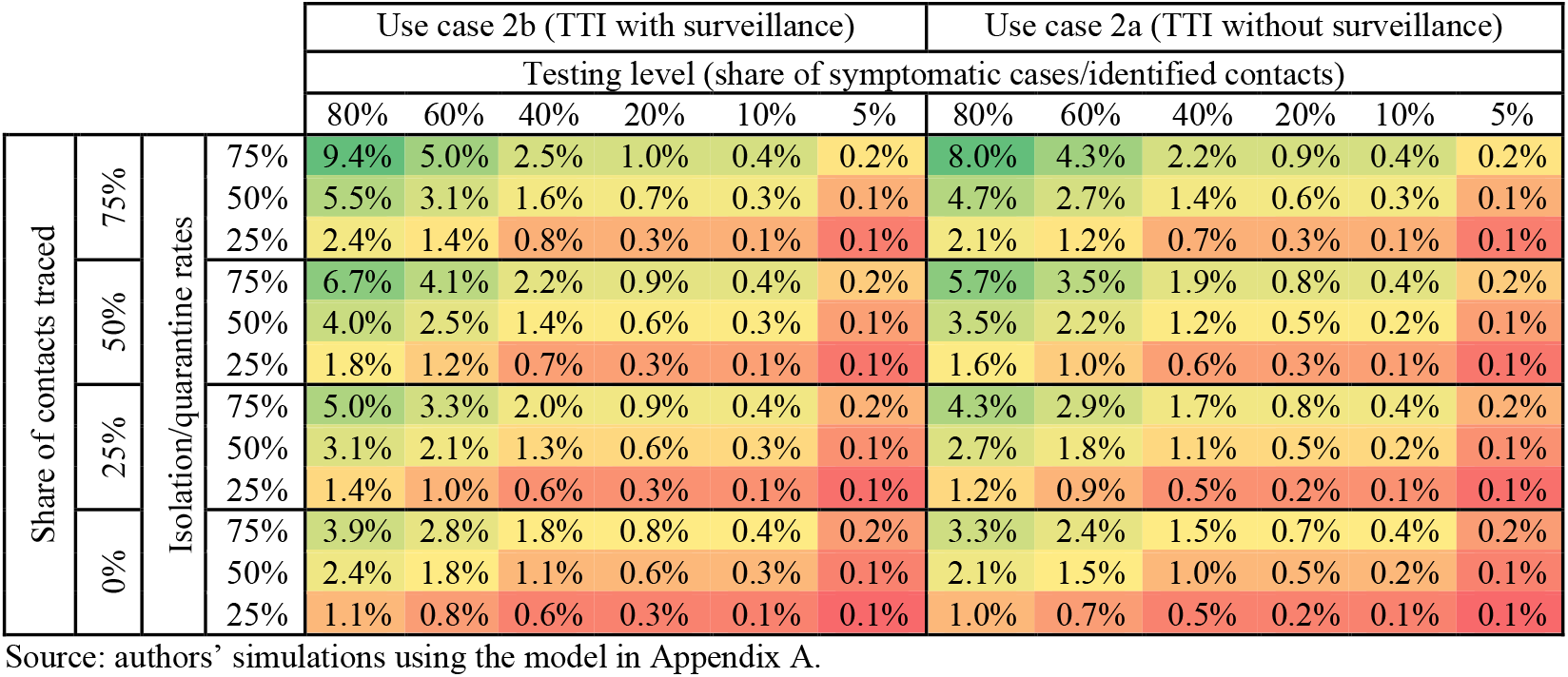
Percentage reduction in peak ICU needs in different TTI scenarios

**Table 3:**
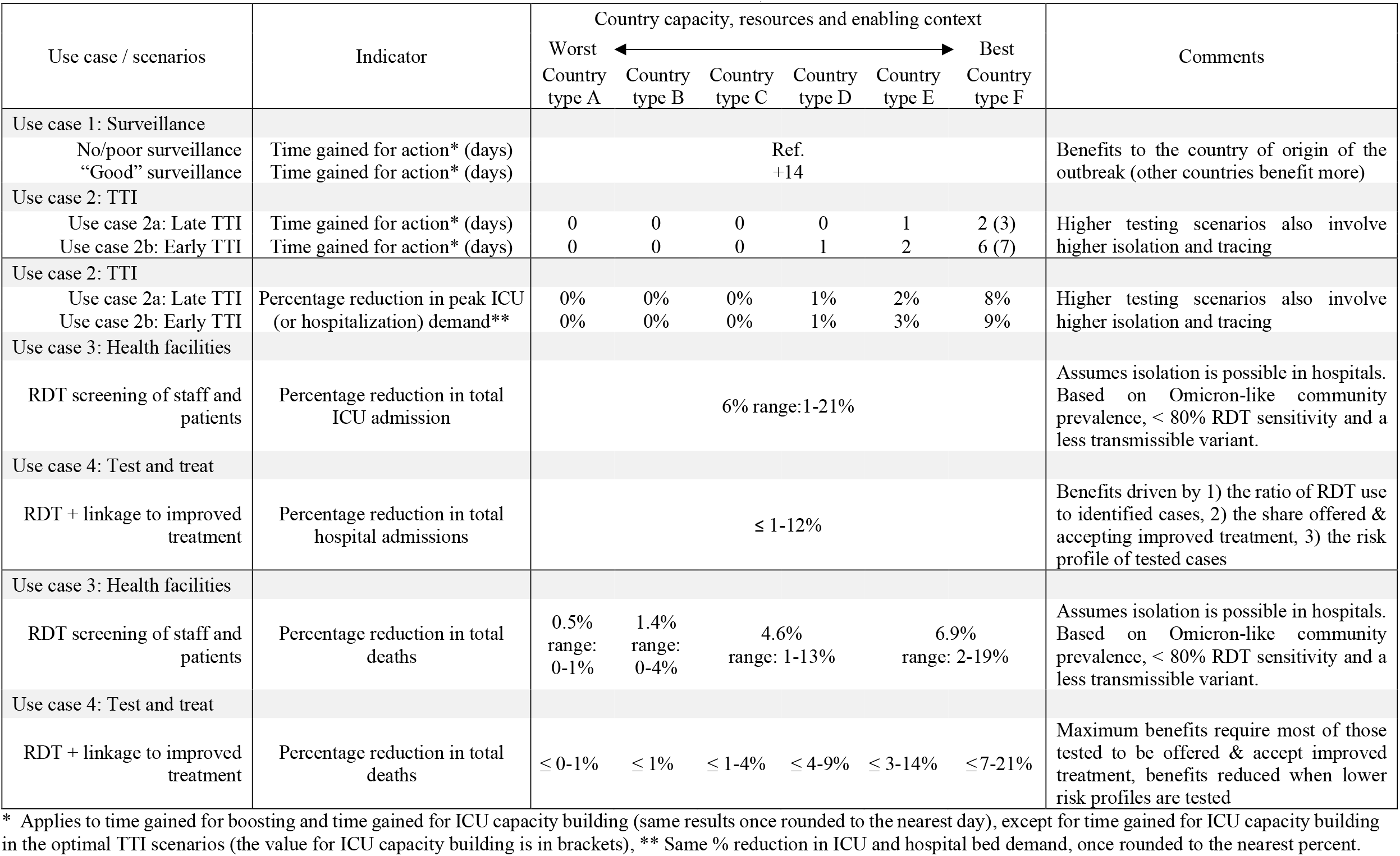
Summary results: impacts of different use cases on time gained, peak health system burden and deaths

Short-term transmission reduction decreases peak needs. We measure that impact through an assessment of changes in “unmet needs” (demand for ICU beds exceeding official capacity). In “optimal” (80% of symptomatic cases tested, 75% traced and 75% isolating) and “high” (60% tested, 50% traced and 50% isolating) scenarios, the reductions in “unmet needs” that can be achieved are at most 5% and 1.3% of total (met and unmet) needs, respectively. Appendix B describes how “unmet needs” evolve with ICU capacity.

Note that combined transmission reduction interventions are more effective than the sum of isolated interventions: intervening one day earlier with a 40% reduction in transmission achieves a gain of 0.69 days for boosting or capacity strengthening, or around 3 times the benefits of 20% transmission reduction (gain of 0.24 days). Similar results are found for the reduction in peak ICU needs.

### Use case 3: Reduction in nosocomial transmission

Using the assumptions described in Methods, the use of RDTs in hospital settings may reduce ICU demand by 5.9% of total demand (median parameters in baseline scenario), with ranges from 1.5-20.6% depending on parameter estimates. The use of RDTs may further reduce in-hospital COVID deaths by around 9% (extreme values: 3-25%). Using death ascertainment scenarios of 5%, 15%, 50% and 75% of total deaths (close to median death ascertainment in low-, lower-middle-, upper-middle- and high-income countries respectively [2]) and assuming this represents a good estimate of the share of deaths happening in hospitals, RDTs to reduce nosocomial COVID may decrease total deaths by 0.5% (0-1%) to 6.9% (2-19%) (Table 3).

### Use case 4: Testing and treatment

We assessed six scenarios detailed in Appendix F corresponding to RDT access of 5% to 80%. The reduction in hospitalization ranges from 1 to 12% if improved treatment is offered to and accepted by those that test positive with RDTs. Meanwhile, the reduction in deaths increases with RDT access and ranges from 0-1% to 7-21% (Table 3).

### Summary table

Table 3 summarizes the results of analyses for baseline outbreak characteristics and test sensitivity for six country “archetypes” going from low-resource/low-capacity contexts (Archetype A) to high-resource/high-capacity contexts (Archetype E and F, F being an “ideal” scenario in which all parameters are optimized). Appendix F summarizes the parameters used in each scenario.

### Sensitivity analysis

Table 4 provides the results of the sensitivity analysis, using the simulation model in Appendix A and formulas in the methodology. Further details are provided below and in Appendix D. β represents the effective contact rate and *s*_*0*_ is the share of the population that is susceptible to the outbreak when it starts (*s*_*0*_=50% in the baseline scenario, 100% represents a highly immune-escaping variant while 34% represents higher baseline immunity). For time gain, the numbers in bracket represent the time available for ICU capacity strengthening.

**Table 4:** Sensitivity analysis results

| Modified parameter | Base case | $\beta$ multiplied by 0.68 | $\beta$ doubled | $s_0 = 34\%$ | $s_0 = 100\%$ | infectious period doubled | latent period halved | RDT sensitivity halved |
| --- | --- | --- | --- | --- | --- | --- | --- | --- |
| r value | r = 5 | r = 3.4 | r = 10 | r = 3.4 | r = 10 | r = 10 | r = 5 | r=5 |
| Use case 1: Surveillance |  |  |  |  |  |  |  |  |
| Time available for boosting (resp. ICU capacity building) with surveillance | 17 (47) | 28 (57) | 8 (37) | 25 (55) | 9 (39) | 14 (46) | 11 (41) | 14 (44) |
| Time available for boosting (resp. ICU capacity building) without surveillance | 3 (33) | 9 (39) | 0 (28) | 7 (36) | 0 (30) | 1 (32) | 0 (30) | 3 (33) |
| Time gained for boosting or ICU capacity building through the use of surveillance | 14 | 19 | 9 | 19 | 9 | 14 | 11 | 11 |
| Use case 2a: TTI* (without surveillance) |  |  |  |  |  |  |  |  |
| Time to boost (resp. build ICU capacity) | 2 (3) | 5 (5) | 1 (1) | 3 (3) | 1 (1) | 1 (1) | 2 (2) | 1 (1) |
| Percentage reduction in peak ICU demand | 8% | 17% | 2% | 16% | 2% | 3% | 7% | 2% |
| Use case 2b: TTI* (with surveillance) |  |  |  |  |  |  |  |  |
| Time to boost (resp. build ICU capacity) | 6 (7) | 12 (13) | 3 (3) | 11 (10) | 3 (3) | 4 (5) | 5 (6) | 2 (2) |
| Percentage reduction in peak ICU demand | 9% | 20% | 2% | 20% | 2% | 4% | 8% | 3% |
| Use case 3: health facilities |  |  |  |  |  |  |  |  |
| Percentage reduction in ICU admissions | Higher $r$ lowers the percentage reduction in nosocomial COVID achievable through screening but increases nosocomial COVID as a share of hospitalized COVID cases. | | | | | | | Lower benefits |
| Use case 4: test and treat |  |  |  |  |  |  |  |  |
| Percentage reduction in hospital admissions | Benefits unchanged if the percentage of cases reached and linked to care does not change. |  |  |  |  |  |  | Benefits halved |
| Percentage reduction in total deaths |  |  |  |  |  |  |  |  |

Source: authors' simulations using the model in Appendix A.
\* The values used for these simulations correspond to an optimal testing, tracing and isolation scenario (country type F or 80% tested, 75% isolating and 75% traced)

Time available for boosting or increasing ICU capacity in the surveillance and no surveillance scenarios reduces for higher effective contact rates β, higher initial shares of susceptible individuals *s*_*0*_ or lower latent periods. Meanwhile, a halving in RDT sensitivity delays outbreak detection with surveillance by 2.8 days.

In use cases 2a and 2b, lower test sensitivity reduces the impact of TTI on transmission reduction for a given level of advance warning, while a higher *r* reduces the impacts of a given level of transmission reduction on time available and hospital burden. This compounds changes in the level of advance warning brought by changes in variant characteristics or test sensitivity. Lower warning reduces the impact of TTI on all outcomes, though for the percentage reduction in peak ICU demand, this effect is minimal.

In use case 3, [30] shows that RDT screening in healthcare settings is less effective (lower percentage reduction in nosocomial COVID prevalence) when community prevalence increases and/or when test sensitivity decreases. Further, transmission reduction in general is more challenging to achieve with more transmissible variants (see use case 2). However, as the advent of Omicron has been associated with increases in nosocomial COVID prevalence [22], a more minor percentage reductions in nosocomial COVID prevalence may still represent a sizeable share of all hospitalized COVID cases.

Finally, the estimates for use case 4 do not directly relate to variant characteristics. However, the benefits of test and treat (even neglecting the harm of false negatives) decrease as test sensitivity decreases. Further, as the outbreak is changing, resistance to existing therapeutics may develop [35], reducing treatment effectiveness.

## DISCUSSION

In optimal scenarios and for median parameters (Omicron-like outbreak and median estimates for contextual and impact parameters), all use cases except TTI without surveillance could allow (depending on the corresponding outcomes) time gains of at least six days and/or reduction in ICU or hospital admissions, peak ICU needs, or deaths over 6%, while TTI without surveillance does not provide much time gain but can still reduce peak ICU needs by 8%. Time gains may allow for better response in terms of ICU capacity building or boosting. Yet, even in the best-case scenario, the boosting response in a median LMIC at the speed at which vaccination was first rolled-out would not exceed 6% of 60+ years old reached. Our results suggest that RDTs alone will not dramatically reduce the burden of COVID-19 in LMICs, but that they may have an important role alongside other interventions that countries are considering such as vaccination, therapeutic drugs, improved healthcare capacity and non-pharmaceutical measures such as improved ventilation and mask wearing.

The ability to achieve results declines when test sensitivity or testing levels are lower as well as when an outbreak is more transmissible or immune escaping. Some use cases are less sensitive to test availability than others, in particular interventions that do not require high testing levels (RDT screening in healthcare settings and surveillance). Further, given the assumptions associated with use case 4 (reaching easier to reach patients first and providing them with high levels of linkage to care), test and treat retains a relatively high potential impact on hospital admissions even with low levels of testing. Our results are coherent overall with the evolution of current guidance, which places decreasing emphasis on the use of RDTs for transmission reduction. They however allow for the comparison of a broad range of use cases, country profiles, variant and test sensitivity scenarios.

While this paper provides information on the impact of different RDT use cases, policy decision-making would also require an understanding of the cost-effectiveness of different options, particularly in settings with limited financial resources. Costs are driven by the number of tests undertaken and the nature of follow-up actions e.g., antiviral treatment, as well as cost savings from reduced healthcare use and improved productivity. They may also depend on the characteristics of the outbreak: for example, a more immune-escaping variant leads to higher numbers of infections over the course of the outbreak, hence achieving a given scenario (e.g., 20% of symptomatic cases tested) may become substantially costlier. Decision-making should also rely on comparisons with interventions beyond RDT use cases, which were not the focus of this paper. Finally, should a highly transmissible, immune-escaping and lethal variant emerge there would be a need to expand the range of interventions considered in this paper beyond strategies to mitigate burden without preventing widespread transmission, potentially reverting back to virus suppression strategies.

There are a number of limitations in this paper: some key assumptions e.g., that hospitals are able to isolate COVID patients may not hold in the least resourced settings. Reaching maximum benefits with test and treat requires optimal linkage to care, which is unlikely in many countries, even in some high-income settings. The negative impacts of false positives or negatives as opposed to no test at all (e.g., unnecessary or delayed treatment) has not been accounted for, but neither has the benefit of being able to rapidly rule out other causes of acute respiratory symptoms/fever that require different treatments (such as bacterial infections and malaria). Further, the variety in outcomes, a necessary consequence of assessing a broad range of interventions, reduces the ability to make comparisons across use cases. Outcomes expressed as a reduction in hospital burden further have to be interpreted with caution, as the meaning of a 5% reduction in hospital burden is not the same in a country in which most severe and critical cases access hospitals vs. a country in which only a minority do. This paper also relies on several simplifications including: uniform test sensitivity, fixed latent, pre-clinical and clinical periods in transmission reduction formulas, and a simplified simulation model. Scenarios were further developed to reflect a “typical” country at a given income level hence do not reflect any specific country characteristics. Modelling to inform detailed policy-making at a country level will require more complex models, country-specific data and involvement of local analysts and policy-makers. Finally, the choice of outcomes leaves out indirect benefits (e.g., how reduced hospital burden may translate into fewer deaths) and a number of additional benefits that are harder to quantify, such as how outbreaks and reducing the number of those affected could also delay the emergence of new variants [36] and, by reducing baseline prevalence, enhance the ability of surveillance systems to identify outbreaks.

## CONCLUSION

The impacts of RDT use cases (surveillance, TTI, RDT screening in health facilities, and test and treat) differ across countries and outbreak types. Large-scale TTI with good surveillance can reduce peak ICU demand and delay the peak of the outbreak by a week, which compounds the benefits of surveillance. The impact of TTI however drops rapidly as the scale of testing and country income level reduce. Oher use cases are somewhat less dependent on large-scale RDT availability. The emergence of new, more transmissible variants, escaping immunity, resistant to existing therapeutics and/or reducing the sensitivity of existing tests, could however decrease the impact of any intervention. Policy decision-making should integrate an assessment of costs and compare both RDT use cases and other potentially relevant interventions accounting for uncertainty on variant characteristics.

## Data Availability

All data produced in the present work are contained in the manuscript

## PATIENT AND PUBLIC INVOLVEMENT

Patients and/or the public were not involved in the design, or conduct, or reporting, or dissemination plans of this research.

