## Appendices for "Is there a role for RDTs as we live with COVID? An assessment of different strategies"

#### APPENDIX A – METHODOLOGICAL SUPPLEMENTS

##### Description of the simulation model and parameters

The model used throughout this paper describes a well-mixed population, with compartments for people who are susceptible (density *s*), exposed (density *e*), never-hospitalized infected (density $i_{nh}$), pre-hospitalized infected (density $i_{ph}$), general ward hospitalized (density $h_g$), pre-ICU hospitalized i.e., hospitalized in the general ward that will ultimately be moved to ICU (density $h_{preICU}$), and ICU (density $h_{ICU}$). We ignore natural deaths and births, as well as disease-induced deaths, given the short timescales over which we are modelling. *H* represents the share of all COVID cases that will need to be hospitalized. All model parameters are detailed in Table A. The model was implemented in R using a time step of 0.01 days for simulations. Outbreaks were modelled using an initial share of infectious individuals equal to 1 per million people. Model equations are as follows:

$$\frac{ds}{dt} = -\beta\, s\,(i_{nh} + i_{ph})$$

$$\frac{de}{dt} = \beta s\,(i_{nh} + i_{ph}) - \gamma_1\, e$$

$$\frac{di_{nh}}{dt} = \gamma_1 (1-h) e - \gamma_2\, i_{nh}$$

$$\frac{di_{ph}}{dt} = \gamma_1 h\, e - \gamma_h\, i_{ph}$$

$$\frac{dh_g}{dt} = \gamma_h\, i_{ph}\,(1 - p_{ICU}) - \gamma_g\, h_g$$

$$\frac{dh_{preICU}}{dt} = \gamma_h\, i_{ph}\, p_{icu} - \gamma_{preICU}\, h_{preICU}$$

$$\frac{dh_{ICU}}{dt} = \gamma_{preICU}\, h_{preICU} - \gamma_{ICU}\, h_{ICU}$$

**Table A1: key model parameters**

| Indicator | Value |
|---|---|
| Effective contact rate β | Calculated based on model parameters for different $R_0$ using the leading eigenvalue method ($R_0$=10 in the baseline scenario hence $\beta$ = 1.7). |
| Share of all cases that are hospitalized *h* | 1%: assumption. *h* depends on each country's age/comorbidity profile, past immunity and variant and was estimated at 0.9% for Omicron in the UK [1]. However, results in this paper do not depend on its exact value given the indicators used for results (% reduction). |
| Mean latent period $1/\gamma_1$ | 4 days [2] |
| Mean duration of infectiousness $1/\gamma_2$ | 6 days – based on 5.6 days for symptomatic cases and 7 days for asymptomatic cases, 35% of cases being asymptomatic [2, 3] |
| Mean delay from onset of clinical symptoms to hospitalization | 7 days [2] |
| Mean delay from infection to hospitalization $1/\gamma_h$ | 2.4 + 7 days (pre-symptomatic period [2] + duration from onset of clinical symptoms to hospitalization) |
| Duration of hospitalization in a non-ICU bed for a severe case $1/\gamma_g$ | 14.6 days [2] |
| Share of hospitalized cases needing ICU | 30% [2] (varies with variant severity. As for *h*, results are not sensitive to the exact value) |
| Duration of hospitalization in a non-ICU bed, for a critical case (assumed to take place before transfer to a ICU) $1/\gamma_{preICU}$ | 6 days [2] |
| Duration of hospitalization in an ICU bed for a critical case $1/\gamma_{ICU}$ | 9.6 days [2] |

#### Summary of scenario characteristics

Table A2 summarizes the characteristics of the scenarios described in Table 3 in the main text. Each scenario corresponds to a country "archetype" defined by a coherent set of characteristics going from a low-capacity, low-resource country (country type A) to a high-capacity, high-resource country (country type E) with country type F corresponding to optimal conditions for all parameters. Justifications for the parameters selected for the different scenarios are provided either in Methods (for short justifications) or in the corresponding appendices (Appendix B for TTI, and Appendix C for testing and treatment). Parameters such as death ascertainment were derived from median conditions in low-income (for country type A), lower-middle-income (for country type B), upper-middle-income (for country types C and D) and high-income (for country types E and F) countries. However, what matters for a country is not its income level but what parameter set is closest to the local conditions for the specific use case. For example, a low-income country with 20% RDT use among symptomatic cases, 25% of people testing positive isolating, and 25% of contacts of positive cases being traced would correspond to Country type C for the purpose of estimating the impact of TTI.

**Table A2: summary of scenario characteristics**

| Use case / scenarios | Country capacity, resource and enabling context<br>Worst ←→ Best<br>Country type A | Country type B | Country type C | Country type D | Country type E | Country type F |
|---|---|---|---|---|---|---|
| Surveillance | | | | | | |
| No/poor surveillance | Outbreak identified when hospital admissions increased | | | | | |
| "Good" surveillance | Outbreak identified through case increases | | | | | |
| TTI | | | | | | |
| Late TTI<br>Early TTI | 5% RDT use*, 25% isolated, 0% traced | 10% RDT use*, 25% isolated, 0% traced | 20% RDT use*, 25% isolated, 25% traced | 40% RDT use*, 50% isolated, 25% traced | 60% RDT use*, 50% isolated, 50% traced | 80% RDT use*, 75% isolated, 75% traced |
| Nosocomial testing | | | | | | |
| RDT screening | RDT screening of patients/staff in addition to routine PCR<br>5% death ascertainment | 15% death ascertainment | 50% death ascertainment | | 75% death ascertainment | |
| Test and treat | | | | | | |
| RDT + linkage to improved treatment | 5% RDT use*, 5% death ascertainment, 15% of severe treated, 0.1-0.3% of cases benefit from optimal PCR + care | 10% RDT use*, 15% death ascertainment, 40% of severe treated, 1-3% of cases benefit from optimal PCR + care | 20% RDT use*<br>50% death ascertainment, 75% of severe treated, 5-15% of cases benefit from optimal PCR + care | 40% RDT use* | 60% RDT use*<br>75% death ascertainment, 90% of severe treated, 15-45% of cases benefit from optimal PCR + care | 80% RDT use* |

* This corresponds to the expected level of RDT use among COVID cases upon symptom onset.

### APPENDIX B – USE CASES 2A AND 2B: TTI

#### Testing and isolation

We assume that all durations (e.g., infectious periods) are fixed. We note: $s$ = test sensitivity, $t$ = percentage of all symptomatic cases tested, $i$ = isolation probability upon a positive test, $\alpha$ = share of asymptomatic cases, $T_{asym}$ = infectious period, $f$ = relative infectivity for asymptomatic cases, $T_{pre}$ = preclinical infectious period, $T_{clin}$ = clinical infectious period, $T_{test}$ = time from symptom onset to test, and $\varphi$ = reduction in transmission after isolation starts. The overall reduction in transmission through testing and isolation of clinical cases (without tracing) is then:

$$Transmission\ reduction\ = t\ s\ i\ \varphi\ \frac{(1-\alpha)\ (T_{clin}-T_{test})}{\left(\alpha * T_{asym} * f + (1-\alpha)\ * \left(T_{pre}+T_{clin}\right)\right)}$$

#### Impact of tracing

We first make a number of simplifying assumptions. Traced contacts may have been exposed at any point between the start of the infective period and the moment the index's test results are known and the index is isolated. The earliest infected contacts were exposed at the beginning of the pre-symptomatic period (2.4 days before symptom onset [2]). We have further assumed that, for RDTs, the average delay between symptom onset and testing is 0.5 days and that test results are immediate, hence contacts are generally infected within 2.9 days of the index's test. We further assume that hard to trace contacts are not routinely traced and that traced contacts are reached within a day, or within 3.9 days of infection. The latent period (between exposure and the beginning of the infective period) has been set at 4 days [2], hence, most contacts will be informed they were exposed before the end of their latent period, and we have assumed that we can neglect secondary transmission from traced contacts taking place before they are informed of their exposed status. We make the following additional simplifying assumptions:

- No policy is set in place to trace the contacts of traced contacts (expected to start isolating before they become infective).
- No system is in place to trace the contacts of cases that did not test or tested negative (these cases are not identified).
- A contact that refuses to test will not isolate, even if they are symptomatic.
- People who do not isolate may inform their contacts, hence may trace but not isolate. They may also isolate but not trace (e.g., if concerned with stigma) or do both or neither.

Table B1 estimates the percentage reduction in onward transmission resulting from index isolation and contact tracing. In this table and future calculations, we use the following notation and assumptions:

- $q$ is the share of all index cases that have been identified as contacts of an infected case.
- $t$ and $t'$ are the shares of symptomatic cases and identified contacts, respectively, that test.
- $s$ and $s'$ are the values of test sensitivity for symptomatic cases and identified contacts.
- $i$ and $i'$ are the probabilities for symptomatic cases and identified contacts to isolate after a positive test.
- $\varphi$ is the reduction in transmission after isolation starts.
- $\alpha$, $T_{asym}$ and $f$ are the share of asymptomatic cases, the duration of their infective period and their relative infectivity, respectively.
- $T_{pre}$ and $T_{clin}$ are the durations of the pre-symptomatic and symptomatic infective periods while $T_{test}$ is the time from the symptom onset to the test.
- $tr$ is the proportion of cases testing positive after developing symptoms that are traced.
- $\beta$ is the effective contact rate described in Appendix A.

In Table B1, each line corresponds to a category of infected individuals (e.g., individuals who know they have been exposed to the disease, decide to test, for whom the test result is positive, and deciding to isolate). The last two columns estimate secondary transmission from that category without testing (no isolation/tracing) and transmission reduction with testing. The values shown in these columns are the product of the share of infected individuals belonging to the category (depending on hypotheses made regarding e.g., the share of people accepting to test), and the level of onward transmission from each individual within the group.

**Table B1: Plausible testing, isolation and tracing scenarios for individuals that are infected with COVID**

| Categories of infected individuals | | | | | | Level of secondary transmission if testing were not available (no isolation/tracing) | Reduction in secondary transmission with testing |
|---|---|---|---|---|---|---|---|
| **Identified contact[1]** | **Symptom status[2]** | **Test[3]** | **Test result[4]** | **Isolation[5]** | **Tracing[6]** | | |
| Yes | Any | Yes | + | Yes | No | $q\ t'\ s'\ i\ \beta\ L$ | $q\ t'\ s'\ i'\ \beta\ L\ \varphi$ |
| Yes | Any | Yes | + | No | No | $q\ t'\ s'\ (1 - i)\ \beta\ L$ | 0 |
| Yes | Any | Yes | - | No | No | $q\ t'\ (1 - s')\ \beta\ L$ | 0 |
| Yes | Any | No | NA | No | No | $q\ (1 - t')\ \beta\ L$ | 0 |
| No | Asymptomatic | No | NA | No | No | $(1 - q)\ \alpha\ \beta\ a\ f$ | 0 |
| No | Symptomatic | Yes | + | Yes | Yes | $(1 - q)\ (1 - \alpha)\ t\ s\ i\ tr\ \beta\ (T_{pre} + T_{clin})$ | $(1 - q)\ (1 - T_{asym})\ t\ s\ i\ tr\ \beta\ (T_{clin} - T_{test})\ \varphi$ |
| No | Symptomatic | Yes | + | Yes | No | $(1 - q)\ (1 - \alpha)\ t\ s\ i\ (1 - tr)\ \beta\ (T_{pre} + T_{clin})$ | $(1 - q)\ (1 - T_{asym})\ t\ s\ i\ (1 - tr)\ \beta\ (T_{clin} - T_{test})\ \varphi$ |
| No | Symptomatic | Yes | + | No | Yes | $(1 - q)\ (1 - \alpha)\ t\ s\ (1 - i)\ tr\ \beta\ (T_{pre} + T_{clin})$ | 0 |
| No | Symptomatic | Yes | + | No | No | $(1 - q)\ (1 - \alpha)\ t\ s\ (1 - i)\ (1 - tr)\ \beta\ (T_{pre} + T_{clin})$ | 0 |
| No | Symptomatic | Yes | - | No | No | $(1 - q)\ (1 - \alpha)\ t\ (1 - s)\ \beta\ (T_{pre} + T_{clin})$ | 0 |
| No | Symptomatic | No | NA | No | No | $(1 - q)\ (1 - \alpha)\ (1 - t)\ \beta\ (T_{pre} + T_{clin})$ | 0 |

[1] Refers to whether the infected individuals are traced contacts or not.
[2] Differentiates infected individuals depending on their symptoms: asymptomatic (never developing symptoms), symptomatic (developing symptoms some time after exposure), any (may or may not develop symptoms).
[3] Refers to whether the infected individuals test or not.
[4] Test result: refers to whether the test result is positive or negative (NA is for individuals that did not test).
[5] Whether the individual decides, upon a positive test result, to isolate or not.
[6] Whether an infected individual traces their contacts (for simplicity, represented as a yes/no answer in the table though some infected individuals will trace part but not all of their contacts).

Considering from now on (for simplicity) that $t = t'$, $s = s'$ and, $i = i'$, and denoting $L = \left(\alpha\ T_{asym}\ f\ +\ (1-\alpha)\left(T_{pre} + T_{clin}\right)\right)$, we have:

$$\%\ reduction\ in\ transmission =\ t\ s\ i\ \ \varphi\ \left(q + (1-q)\frac{(T_{clin} - T_{test})}{L}\ \right)$$

$$q = \frac{t\ s\ tr\ (1-\alpha)\left(T_{pre} + T_{clin}\right)}{L - t\ s\ tr\ (1-\alpha)\left(T_{pre} + T_{clin}\right)}$$

$$\%\ reduction\ in\ transmission =\ t\ s\ i\ \ \varphi\frac{T_{clin} - T_{test}}{L}\left(1 +\ t\ s\ tr\frac{\left(L - (1-\alpha)(T_{clin} - T_{test})\right)\left(T_{pre} + T_{clin}\right)}{\left(L - t\ s\ tr\ (1-\alpha)\left(T_{pre} + T_{clin}\right)\right)\ (T_{clin} - T_{test})}\right)$$

#### Transmission reduction scenarios

Table B2 details the reduction in transmission associated with different TTI scenarios.

**Table B2: reduction in transmission from TTI, according to the share of cases/contacts tested, traced and isolating/quarantining upon learning test results**

| Share of contacts traced | | Isolation/quarantine rates | Share of clinical cases & traced contacts tested: 80% | 60% | 40% | 20% | 10% | 5% |
|---|---|---|---|---|---|---|---|---|
| | 75% | 75% | 24.2% | 15.0% | 8.4% | 3.6% | 1.6% | 0.8% |
| | | 50% | 16.1% | 10.0% | 5.6% | 2.4% | 1.1% | 0.5% |
| | | 25% | 8.1% | 5.0% | 2.8% | 1.2% | 0.5% | 0.3% |
| | 50% | 75% | 18.9% | 12.6% | 7.5% | 3.4% | 1.6% | 0.8% |
| | | 50% | 12.6% | 8.4% | 5.0% | 2.2% | 1.1% | 0.5% |
| | | 25% | 6.3% | 4.2% | 2.5% | 1.1% | 0.5% | 0.3% |
| | 25% | 75% | 15.0% | 10.7% | 6.7% | 3.2% | 1.6% | 0.8% |
| | | 50% | 10.0% | 7.1% | 4.5% | 2.1% | 1.0% | 0.5% |
| | | 25% | 5.0% | 3.6% | 2.2% | 1.1% | 0.5% | 0.3% |
| | 0% | 75% | 12.1% | 9.1% | 6.1% | 3.0% | 1.5% | 0.8% |
| | | 50% | 8.1% | 6.1% | 4.0% | 2.0% | 1.0% | 0.5% |
| | | 25% | 4.0% | 3.0% | 2.0% | 1.0% | 0.5% | 0.3% |

Source: authors' calculations based on the formula in Appendix B.

#### Time gained for ICU capacity strengthening

Table B3 provides the time gained through different TTI scenarios, between the start of the intervention and the moment half of ICU bed-days associated with the outbreak have already been used. Results are very similar to those of Table 1 in the main text, which focused on time gained for boosting. Given the results found in Use case 1 (47.2 days between outbreak detection with surveillance and the moment half of ICU bed days have been used, in the absence of TTI, the total time available for ICU-specific capacity building with surveillance is the time gained for ICU capacity strengthening in Table B3, added to 47.2 days e.g., in the best TTI scenario would be 6.8 + 47.2 = 54.0 days.

**Table B3: Time gained for ICU capacity strengthening through different TTI scenarios**

| | | | Benefits of TTI in the presence of surveillance (use case 2b) | | | | | | Benefits of TTI in the absence of surveillance (use case 2a) | | | | | |
|---|---|---|---|---|---|---|---|---|---|---|---|---|---|---|
| | | | Testing level (share of symptomatic cases/identified contacts) | | | | | | | | | | | |
| Share of contacts traced | | Isolation/quarantine rates | 80% | 60% | 40% | 20% | 10% | 5% | 80% | 60% | 40% | 20% | 10% | 5% |
| | 75% | 75% | 6.79 | 3.67 | 1.88 | 0.75 | 0.34 | 0.16 | 2.52 | 1.38 | 0.71 | 0.29 | 0.13 | 0.06 |
| | | 50% | 4.00 | 2.29 | 1.21 | 0.49 | 0.23 | 0.11 | 1.50 | 0.86 | 0.46 | 0.19 | 0.09 | 0.04 |
| | | 25% | 1.79 | 1.07 | 0.59 | 0.25 | 0.11 | 0.06 | 0.68 | 0.41 | 0.22 | 0.10 | 0.05 | 0.02 |
| | 50% | 75% | 4.88 | 2.98 | 1.66 | 0.71 | 0.33 | 0.16 | 1.82 | 1.12 | 0.63 | 0.27 | 0.13 | 0.06 |
| | | 50% | 2.97 | 1.88 | 1.07 | 0.47 | 0.22 | 0.11 | 1.12 | 0.71 | 0.41 | 0.18 | 0.09 | 0.04 |
| | | 25% | 1.37 | 0.89 | 0.52 | 0.23 | 0.11 | 0.06 | 0.52 | 0.34 | 0.20 | 0.09 | 0.04 | 0.02 |
| | 25% | 75% | 3.68 | 2.46 | 1.48 | 0.67 | 0.32 | 0.16 | 1.38 | 0.93 | 0.56 | 0.26 | 0.12 | 0.06 |
| | | 50% | 2.29 | 1.57 | 0.96 | 0.44 | 0.21 | 0.11 | 0.86 | 0.59 | 0.36 | 0.17 | 0.08 | 0.04 |
| | | 25% | 1.07 | 0.75 | 0.47 | 0.22 | 0.11 | 0.06 | 0.41 | 0.29 | 0.18 | 0.09 | 0.04 | 0.02 |
| | 0% | 75% | 2.84 | 2.05 | 1.32 | 0.64 | 0.31 | 0.16 | 1.07 | 0.77 | 0.50 | 0.24 | 0.12 | 0.06 |
| | | 50% | 1.80 | 1.32 | 0.86 | 0.42 | 0.21 | 0.11 | 0.68 | 0.50 | 0.33 | 0.16 | 0.08 | 0.04 |
| | | 25% | 0.86 | 0.64 | 0.42 | 0.21 | 0.11 | 0.05 | 0.33 | 0.24 | 0.16 | 0.08 | 0.04 | 0.02 |

Source: authors' simulations using the model in Appendix A

#### Share of 60+ that can be given vaccine boosters during the time gained for boosting

Table B4 details the benefit, in terms of boosting, of different scenarios. The table was computed by:

1) For each country, translating the days available for boosting in each scenario into a number of individuals that could be boosted. For example, for the optimal scenario which gives 23.71 days for boosting, we computed, for the "initial speed", the number of individuals boosted in the country 23.71 days after the start of the vaccination campaign defined as the first day with non-zero numbers of individuals recorded as vaccinated. For the "speed once 1% of the population had been vaccinated", we computed vaccinations per day in the first 14 days after 1% of the population had been vaccinated, then multiplied this number by 23.71.
2) Translating numbers boosted into a share of the 60+ years old population in each country, using World Population Prospects [4] data.
3) Taking the median, 25% and 75% percentiles of the share of 60+ years old boosted for all countries with data in each income range.

A similar computation (not shown below) was undertaken for 80+ years old.

**Table B4: Share of 60+ years old that can be protected through COVID boosters in the time available, by intervention scenario**

| Intervention scenario | | | | Share of 60+ years old boosted (median and IQR) | | | |
|---|---|---|---|---|---|---|---|
| Warning | % tested - traced – isolating | Days to boost | Boosting speed | High income | Upper middle income | Lower middle income | Low income |
| Early | 80%-75%-75% | 23.71 | Initial | 8% [4%,17%] | 5% [2%,19%] | 6% [2%,20%] | 6% [1%,10%] |
| Early | 60%-50%-50% | 19.07 | Initial | 5% [2%,14%] | 3% [1%,12%] | 5% [2%,15%] | 4% [1%,9%] |
| Early | 20%-25%-25% | 17.49 | Initial | 5% [2%,13%] | 3% [1%,10%] | 4% [1%,13%] | 4% [1%,8%] |
| Early | 0%-0%-0% | 17.28 | Initial | 5% [2%,13%] | 3% [1%,9%] | 4% [1%,12%] | 4% [1%,8%] |
| Late | 80%-75%-75% | 5.36 | Initial | 1% [0%,4%] | 1% [0%,2%] | 1% [0%,4%] | 1% [0%,3%] |
| Late | 60%-50%-50% | 3.87 | Initial | 1% [0%,3%] | 0% [0%,2%] | 1% [0%,3%] | 0% [0%,2%] |
| Late | 20%-25%-25% | 3.35 | Initial | 1% [0%,2%] | 0% [0%,1%] | 1% [0%,2%] | 0% [0%,2%] |
| Late | 0%-0%-0% | 3.28 | Initial | 1% [0%,3%] | 0% [0%,2%] | 1% [0%,3%] | 0% [0%,2%] |
| Early | 80%-75%-75% | 23.71 | 1% vacc. | 13% [10%,26%] | 18% [10%,32%] | 22% [12%,41%] | 9% [4%,15%] |
| Early | 60%-50%-50% | 19.07 | 1% vacc. | 11% [8%,21%] | 14% [8%,25%] | 18% [10%,33%] | 8% [3%,12%] |
| Early | 20%-25%-25% | 17.49 | 1% vacc. | 10% [8%,19%] | 13% [8%,23%] | 16% [9%,30%] | 7% [3%,11%] |
| Early | 0%-0%-0% | 17.28 | 1% vacc. | 10% [7%,19%] | 13% [8%,23%] | 16% [9%,30%] | 7% [3%,11%] |
| Late | 80%-75%-75% | 5.36 | 1% vacc. | 3% [2%,6%] | 4% [2%,7%] | 5% [3%,9%] | 2% [1%,3%] |
| Late | 60%-50%-50% | 3.87 | 1% vacc. | 2% [2%,4%] | 3% [2%,5%] | 4% [2%,7%] | 2% [1%,3%] |
| Late | 20%-25%-25% | 3.35 | 1% vacc. | 2% [1%,4%] | 2% [1%,4%] | 3% [2%,6%] | 1% [1%,2%] |
| Late | 0%-0%-0% | 3.28 | 1% vacc. | 2% [2%,4%] | 3% [2%,5%] | 3% [2%,6%] | 1% [1%,2%] |

Source: authors' calculations using [5]'s vaccination dataset, [4]'s population data and the World Bank's country classification [6].

#### Unmet ICU needs

"Unmet needs" were defined as bed-day needs above a certain level designated as "maximum capacity". These correspond to the area of the curve corresponding to bed-day needs for each day of the outbreak that is above the "maximum capacity" level. This value depends on the level constituting "maximum capacity". Figure B1 illustrates the relationship between transmission reduction through TTI (initiated once an outbreak is detected and stopping once infection rates decrease below the level of detection) and unmet ICU bed-day needs. Unmet needs correspond to the area above the line indicated as the "capacity level", and total needs to all the area under the curve. In the left panel, capacity is high though insufficient to meet all needs, while in the right panel, it is very low. In an extreme case (zero ICU capacity), the whole curve would be above the capacity level, and all ICU bed-days needed would be "unmet".

**Figure B1: Relation between capacity level and unmet need**

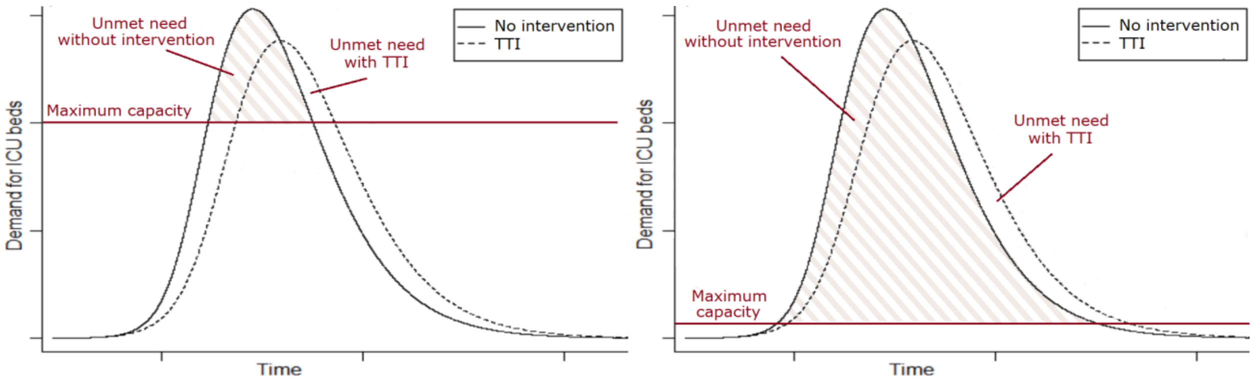


Figure B2 describes the numerical relationship between transmission reduction through TTI and reduction in unmet needs, expressed either as a share of total needs (panel A) or unmet needs (panel B). Maximum capacity has been expressed as a percentage of maximum needs (the maximum daily number of bed-days needed over the course of the outbreak). 0% means no capacity at all and 100% meaning that capacity are at or exceed maximum needs. Two curves are provided in each panel, for two transmission reduction scenarios. Note that unmet needs are reduced through: 1) a "flattening" of the curve when transmission is reduced, meaning that demand for ICU bed-days is more spread out hence strains ICU capacity less 2) a reduction in total demand (which is illustrated by the reduction in unmet needs when there is no capacity).

**Figure B2: reduction in unmet ICU needs as a share of total needs (panel A) and unmet needs (panel B) for different TTI scenarios**

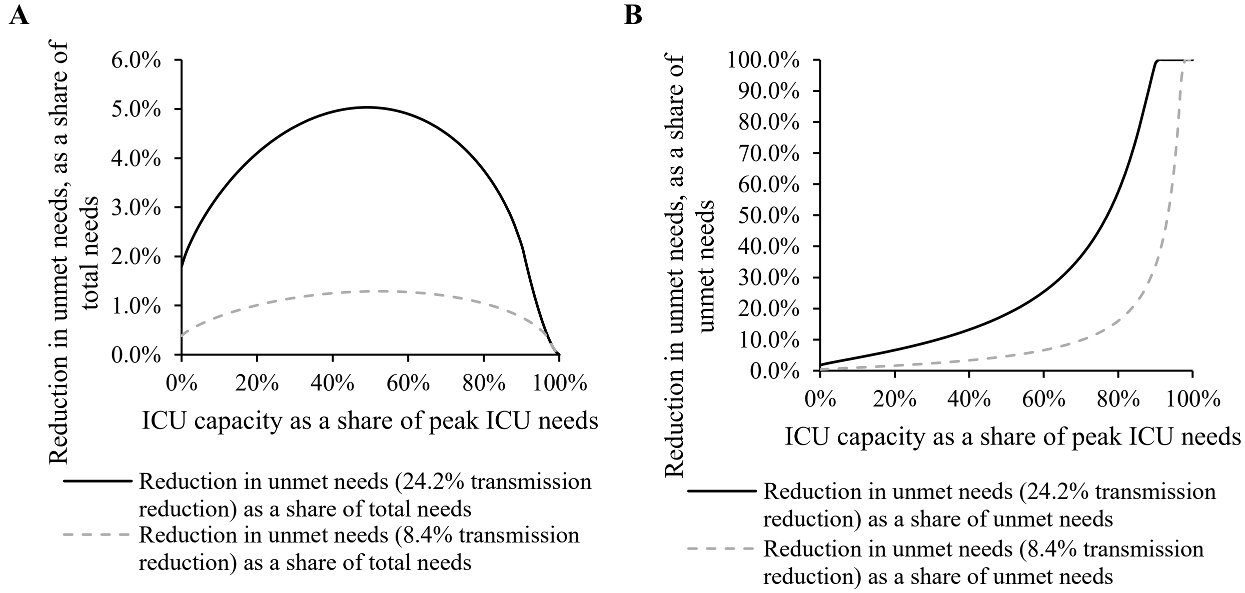


Source: authors' simulations using the model in Appendix A

#### Rationale for scenario assumptions

The scenarios presented in Table 3 and detailed in Table A2 represent a series of country archetypes at different resource and capacity levels., from type A (patterned on low-income settings), type B (lower-middle-income), types C and D (upper-middle-income), type E (high-income) and type F (high-income with optimal parameters). Lower-resource/capacity archetypes have lower RDT availability, tracing capacity, and ability to isolate for individuals testing positive. We further assume that RDTs can lead to a higher relative increase in case identification levels in low-resource settings, where current levels of identification are low, than in higher-resource settings.

The levels of testing selected for each country archetype are: 5% for country type A, (>10 times the 2021 median case identification levels in low-income countries, as per [7] and using a share of asymptomatic cases equal to 35% [3]); 10% for country type B (twice the 2021 median case identification levels in lower-middle income countries); 20% and 40% for country types C and D (once and twice the 2021 median level of case identification in upper-middle-income countries); and 60% and 80% for country types E and F (one and 1.5 times the 2021 median case identification level in high-income countries).

Assumptions for the levels of isolation for each country archetype are based on isolation levels for different types of workers. Reported compliance with isolation ranged from 50%-80% [8, 9] in the UK, while fewer than 20% of informal workers in Mexico (who typically do not have access to paid sick leave and cannot afford to lose income) reported preferring to stop working over getting infected [10] (even fewer may accept to stop working to curb disease transmission). For scenario setting, we assumed that 80% and 0% of workers with and without access to paid sick leave isolate, respectively.

Table B5 reflects workers not covered by paid sick leave provision, either because the country does not have any such provision (all workers are excluded) or because self-employed workers are excluded. Not considered in Table B5 are local paid sick leave provisions in decentralized countries with no national policy, delays to access paid sick leave and levels of pay, and possible exclusion of part-time workers from sick leave policies. Further, the table focuses only on employed workers. Country types A and B are patterned on low- and lower-middle-income countries, where 68% of workers are excluded from paid sick leave provisions. If 80% of those with paid sick leave isolate while none of those without do, then around a quarter of workers would isolate upon testing position – this is the assumption used for country types A and B. Other country types are patterned on upper-middle- and high-income countries. If a third of workers are excluded from sick leave provision and 80% of those with paid sick leave isolate while none of those that are excluded do, around half of infected workers would isolate, a level used in country Types D and E. For type C, we proposed a more modest assumption (lower-performing upper-middle-income context), with 25% isolating. Type F was designed as an optimal scenario, and we assumed 75% of cases would isolate.

**Table B5**: **Paid sick leave provisions: countries and workers covered**

| **Indicator** | **Low income** | **Lower middle income** | **Upper middle income** | **High income** | **LMICs** |
|---|---|---|---|---|---|
| % countries with laws that do not provide sick leave for either all workers or self-workers | 88% | 74% | 45% | 36% | 65% |
| Average share of employed workers in income range excluded from paid sick leave provisions (population-weighted) | 68% | 68% | 32% | 35% | 51% |

Source: data on sick leave provisions for all vs. self-employed workers from © WORLD Policy Analysis Center [42] and data on self-employed workers from ILO [43]

The last assumption relevant to scenario-setting for the TTI use case is the level of tracing. The shares of contacts reached by contact tracers in the UK was close to 50% [11], which was chosen as the share of contacts traced for type E. Type F was built to represent an optimal scenario, and we assumed that 75% of contacts were traced. It was further assumed that for types A and B (patterned on a typical low-income and lower-middle income country), there was insufficient capacity to invest in contact tracing (0% traced). Finally, for types C and D (patterned on upper-middle-income country), an intermediate scenario was chosen (25% traced). There is a broad variety in actual country situations, hence these parameters were selected only for the purpose of filling the summary table in the main text (Table 3) but a larger variety of parameter sets are proposed in the rest of the paper.

### APPENDIX C – USE CASE 4: TESTING AND TREATMENT

Representing the impact of testing associated with referral for improved treatment can be challenging given the variety in plausible scenarios. Figure C highlights how the benefits of treatment change with increasing RDT access:

- We assume that, at the beginning, RDTs tend to reach advantaged populations having good access to PCR tests with low turnaround time and good care. Because these individuals already have access to good diagnosis, RDTs will only save a few hours, which are not critical for early treatment [12].
- We assume that RDTs then reach individuals that have some access to care and limited access to testing i.e., no access to PCR or access to PCR with high turnaround time when they have mild disease, and access to care only if they develop severe or critical symptoms (at which point they will be hospitalized). For these individuals, RDT tests, by providing an early diagnosis, may help them access improved care such as antivirals if available. The shape of the curve in that zone is concave, as larger-scale testing tends to be associated with a lower risk/severity level of those tested.
- At the end of the curve, we assume that the last individuals to be offered RDTs as these are scaled-up are people that currently do not access care even if they have severe symptoms. For these individuals, we assume that, until barriers to linkage to care are resolved, RDTs will not prompt improved care.

**Figure C: Change in treatment benefits with increasing RDT access**

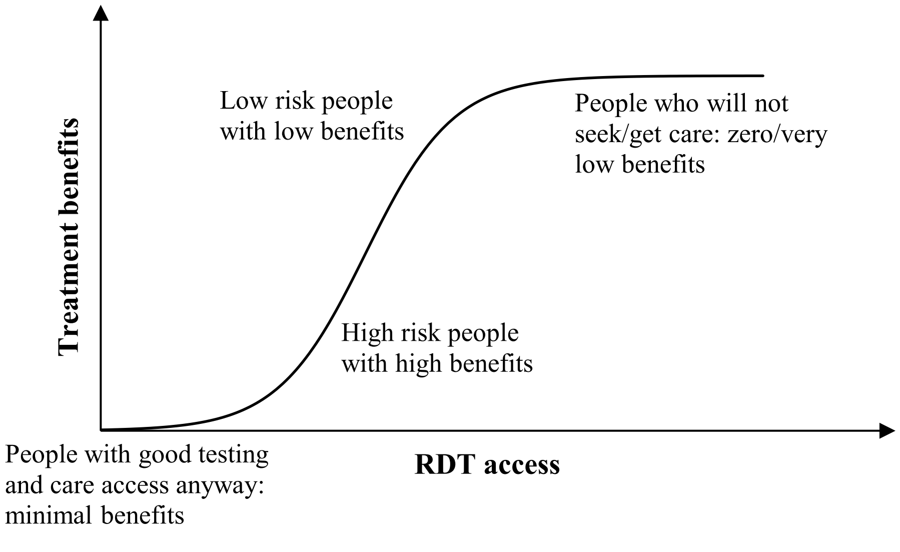


As new therapeutics, research and variants appear, the impact of plausible therapeutics and the target groups that may benefit from them will change. To get an order of magnitude of the impact of treatment when provided, we use the example of the highest performing antiviral as of August 2022 [12, 13], Nirmatrelvir-ritonavir. Given within 3 days of symptom onset, it lowers hospitalization risk to 0.9% from 6.0%, and none of the trial's treated cases died (OR: 0.004 [0-0.67]) [12, 14-16]. However, this is highly uncertain, and effectiveness may differ with new drugs, variants and population profiles. In particular, most of the world population now has some immunity through either vaccination or past infection. It is unlikely that LMIC clinics will be able to triage patients by prior infection status, while the drugs were trialed on unvaccinated, never-infected cases [14, 17, 18]. The results of clinical trials for vaccinated patients with at least one risk factor showed a non-significant reduction in hospitalization or death risk of 57% (from 1.9% to 0.8%) [19]. Further, LMIC high-risk populations are likely to be younger overall than in higher-income countries, and a recent preprint on nirmatrelvir therapy suggests that younger high-risk patients (below 64 years old) may not derive significant benefits from treatment (hospitalization hazard ratio = 0.78) [20]. For the purpose of this paper, we assumed that hospitalization risk for high-risk patients without early treatment is at most 6%, reflecting risks in the trial's placebo group, and at least 1.5%, or a quarter of the original risk, in keeping with realized risk reductions for 60-79 years old (an "at risk" group) in England during the Omicron wave, at a time when vaccination was already widespread [1] (Delta dominated during trials [15, 18]). For treated cases, we use 0.9%, the value found in trials for treated high-risk, unvaccinated never-infected patients and close to the value found for treated, high-risk, vaccinated patients [12, 14-16, 19].

Estimating treatment impact also requires assumptions regarding the share of high-risk cases among hospitalized COVID cases and hospital deaths. High-risk is defined as being 60+ years old or having comorbidities increasing COVID risk. Our assumptions that 25% of hospitalized patients and 50% of those that die are "high-risk" are based on figures for Omicron in South Africa. In this context, 13% of hospitalized Omicron patients were 60+ [21] and 12-23% had comorbidities [21, 22]. Estimating how generalizable these estimates are to other countries is difficult, as the share of older people in the population, the proportion of comorbidities within a given age group, and relative likelihood of hospital access for these groups all change with a country's income level, but in multiple directions.

If $o$ = share of cases benefiting from rapid PCR and good treatment options even in the absence of RDTs (most advantaged segment of the population), $T$ = share of cases that have access to at least some level of care in case of severe disease, even in the absence of RDTs, $t$ = share of cases accessing RDTs thanks to the intervention, $s$ = RDT test sensitivity, $r_h$ = share of high-risk individuals among hospitalized patients (25%), $r_d$ = share of high-risk patients among hospital deaths (50%), $x_h$ = reduction in hospitalization for high-risk severe cases (40-85%) thanks to treatment and $x_d$ = reduction in deaths (67-100%) for these patients, then:

$$Reduction\ in\ hospitalizations \leq s\ \frac{t-o}{T}\ x_h\ r_h$$

$$Reduction\ in\ deaths \leq s\ \frac{t-o}{T}\ x\ r_d * death\ ascertainment$$

The formula involves a number of parameters that have to be defined for each scenario.

- Death ascertainment has been set at 5%, 15%, 50% and 75% for scenarios associated with low- (country type A), lower-middle- (country type B), upper-middle- (country types C and D) and high-income (country types E and F) countries respectively, in line with data in [7].
- We obtained a rough estimate of the share $T$ of cases that have access to some treatment (prior to RDT scale-up), at least if they become severe, using death ascertainment, a case-fatality ratios in hospital around 30% [22, 23] and the assumption that never treated severe cases die. From these

we derive the share of severe cases having some access to identification and treatment in country types A (15%), B (40%), C and D (75%), and E and F (90%) respectively.

- For our scenarios, we have assumed that the share *o* of patients that will not derive benefits from RDT scale-up because they already have access to PCR with short turnaround time and good treatment options increases with case ascertainment. We have set it at 0.1-0.3%, 1-3%, 5-15% and 15-45% in low-, lower-middle-, upper-middle- and high-income scenarios respectively, based on estimates regarding the share of country infections detected in 2021 [24], accounting for asymptomatic infections, and assuming that between 25 and 75% of cases that were identified in 2021 had access to a PCR test with short turnaround time.
- Finally, the scenario is optimal when all high-risk cases that test positive with RDTs (i.e., 80% of cases given test sensitivity) access early/improved care, hence the "≤" in the formula.

Note that, so far, promises of subsidized access to antivirals in LMICs [25-27] and orders of antiviral treatment courses in the US or the UK [28-31] amount to less than 10% and around 30% of their 60+ population, respectively [4], itself only a fraction of high-risk individuals.

### APPENDIX D – COMPLEMENTS TO SENSITIVITY ANALYSIS

The sensitivity analysis in the main body of the paper was developed by simulating outcomes for different outbreak characteristics and test sensitivity. However, it focused only on the way in which results for country type F may evolve with some of the plausible changes in outbreak and test parameters. This section complements that analysis by providing a non-exhaustive set of mathematical formulas for parameters that can formulated in a simple enough manner, for the reader to be able to rapidly adjust the figures discussed in this paper for other country types, different basic reproductive numbers, latent and infective periods, and different abilities to escape immunity (which can affect the value of $s_0$: initially susceptible individual).

#### Use case 1

##### Linear regime

If we neglect hospitalization, we can simplify the differential equations in Appendix A into:

$$(1) \qquad \frac{ds}{dt} = -\beta\, s\, i$$

$$(2) \qquad \frac{de}{dt} = \beta s\, i - \gamma_1\, e$$

$$(3) \qquad \frac{di}{dt} = \gamma_1\, e - \gamma_2\, i$$

As long as cumulative infections are low compared to the initial number of susceptible individuals $s_0$, we can assume that $s = s_0$, transforming equations (2) and (3) into a set of linear equations. These have the following solution, for $i = i_0$ and $e = 0$ at $t = 0$:

$$(4a) \qquad i(t) = -\frac{i_0}{2u}\left((1-\rho-u)e^{\gamma_2\lambda_+ t} - (1-\rho+u)e^{\gamma_2\lambda_- t}\right)$$

$$(4b) \qquad e(t) = \frac{i_0\, r}{u}\left(e^{\gamma_2\lambda_+ t} - e^{\gamma_2\lambda_- t}\right)$$

With $r = \frac{\beta}{\gamma_2} * s_0$ , $\rho = \frac{\gamma_1}{\gamma_2}$ , $u = \sqrt{(\rho-1)^2 + 4\,\rho\, r}$ and $\lambda_+(r) = \frac{-1-\rho+u}{2}$, $\lambda_-(r) = \frac{-1-\rho-u}{2}$

Cumulative infections are low as compared to the initial share of susceptible individuals as long as $\gamma_2 \int_0^t i(\tau)d\tau \ll s_0$. This formula is equivalent to $t \ll \frac{1}{\gamma_2\lambda_+} \ln\left(\frac{2\, s_0 u\, \lambda_+}{i_0\,(\rho+u-1)}\right)$. Further, for $e^{-\gamma_2 ut} \ll 1$, the first term dominates in equations (4a) and (4b). With the values chosen in the baseline scenario and with the condition that the neglected terms in the equations are no more than 10% of the total, then the approximation is valid for 2.5 days < t < 56.5 days.

As long as these conditions are met, we can use the following:

$$(5a) \quad i(t) = -\frac{i_0}{2u}(1-\rho-u)\, e^{\gamma_2\lambda_+ t}$$

$$(5b) \quad e(t) = \frac{i_0\, r}{u}\, e^{\gamma_2\lambda_+ t}$$

#### TIME FROM OUTBREAK DETECTION TO PEAK INFECTIONS

If we assume that a new outbreak is detected when the prevalence of the new variant exceeds a level $i_d$ (e.g., when it is sufficiently above a "background level" of infection corresponding to an earlier variant), then we can use equation (5) to compute the time $t_d$ from the beginning of the outbreak to its detection in a surveillance scenario:

$$(6)\ t_d = \frac{1}{\gamma_2\lambda_+} \ln\left(\frac{-2\, u\, i_d}{i_0(1-\ \rho-u)}\right)$$

[32] provides a mathematical approximation to the total time to outbreak peak $t_{peak}$ for an outbreak starting with a fully susceptible population ($s_0 = 1$). Adapted to account for other values of $s_0$, the formula becomes:

$$(7)\ t_{peak} = \frac{1}{\gamma_2\lambda_+} \ln\left(\frac{\left(1-\frac{1}{r}\right)^2 z_\infty s_0}{\left(z_\infty - 1 + \frac{1}{r}\right) i_0} \frac{\sqrt{(\rho^2+(1+\lambda_+)^2)(\rho^2+(1+\lambda_-)^2)}}{\lambda_-(1+\lambda_-)}\right)$$

$z_\infty$ is the share of those initially susceptible that are ultimately infected through the outbreak. This value is well approximated by

$$(8)\ z_\infty = 1 - e^{-r}$$

For initial infection rates $i_0$ of the magnitude of those used in our simulations (1 per million inhabitants), $i_0$ dominates in (7a), and the dominant term in the equation is:

$$(9)\ t_{peak} \cong -\frac{\ln(i_0)}{\gamma_2\lambda_+}$$

Combining (7) with (6), we can compute the total time from outbreak detection to peak infections. While the logarithm term includes elements that vary with r and other outbreak parameters, overall, the variation in the time from outbreak detection to peak infections is inversely proportional to $\gamma_2\lambda_+$, as below:

$$(10)\ t_{peak} - t_d \sim \frac{Constant}{\gamma_2\lambda_+}$$

Finally, for values of $r \geq 2$ and $\rho$ between 1/3 and 3, we use the approximations: $u \cong \sqrt{4\rho r}$ and $\sqrt{\rho} + \frac{1}{\sqrt{\rho}} \cong 2$ and the time from the beginning of the outbreak to its peak, and from its detection to its peak can be approximated by:

$$(11)\ \ t_{peak} \cong -\frac{\ln(i_0)}{\sqrt{\gamma_1\gamma_2}\,(\sqrt{r}-1)}$$

$$(12)\ \ t_{peak} - t_d \sim \frac{Constant}{\sqrt{\gamma_1\gamma_2}\,(\sqrt{r}-1)}$$

#### Use case 2a and 2b: Impact of TTI on time available for boosting and peak infections

We use the solution to equations (1) to (3) to compute the impact of a reduction in transmission ε on the timing of peak infection rates.

We seek the values of $t_i$ and $t_e$ for which $i(\tau, r) = i\big(\tau + t_i, r(1-\epsilon)\big)$ and $e(\tau, r) = e\big(\tau + t_e, r(1-\epsilon)\big)$, respectively, during the linear period. We find that, in first approximation:

$$(13)\qquad t_i = t_e = \frac{\lambda_+(r) - \lambda_+\big(r(1-\epsilon)\big)}{\lambda_+\big(r(1-\epsilon)\big)}\,\tau$$

$t_i$ = $t_e$ represent the delay in the curve of infected individuals driven by a percentage reduction in transmission ε taking place over a period τ, noting that, with the values chosen in our base scenario, the approximation in (13) is valid for 2.5 days < t < 56.5 days, or almost the entire period between the start of the outbreak and its peak (see linear regime section).

For values of $r \geq 2$, $\rho$ between 1/3 and 3 and a reduction in transmission ε up to 50%, (13) can further be approximated, using $u \cong \sqrt{4\rho r}$ and $\sqrt{\rho} + \frac{1}{\sqrt{\rho}} \cong 2$, into formula (14), which is linear in $\tau$ (the point at which transmission reduction starts), independent of $\rho$ and decreases with $r$:

$$(14)\qquad \frac{\sqrt{r} - \sqrt{r\,(1-\epsilon)}}{(\sqrt{r\,(1-\epsilon)} - 1)}\,\tau$$

The slope provided in (14) decreases with $r$ and tends toward $\varepsilon/2$ for large $r$ and small $\varepsilon$. Simulations show a near-perfect agreement (less than 1% difference) between the theoretical slope in (13) and the slope found in simulations (Figure D1) (agreement with (14) is also good). As outbreak characteristics change, the impact of TTI is affected, on the one hand, by changes in how $\tau$ days of advance warning affect the outbreak (as per (13) and (14)) and by changes in the time available for surveillance. Figure D1 also shows how the timing of interventions affects reduction in peak ICU demand: as long as interventions do not start too close to the peak of the outbreak, the impact of the timing of interventions on peak demand reduction is limited.

**Figure D1: impact of the timing of TTI interventions on time available for boosting and peak ICU needs**

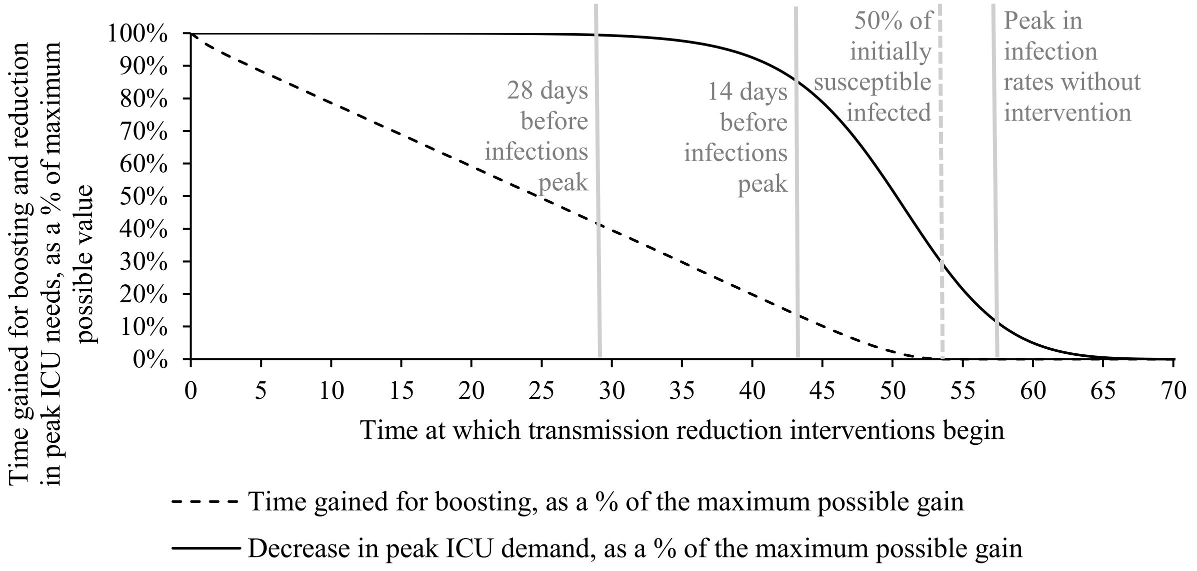


Source: authors' simulations using the model in Appendix A

Peak infection rates computed in [32] are given for an initial share susceptible individuals $s_0$ = 100%, but the formula can easily be adapted for other values of $s_0$, as follows:

$$(15) \qquad Peak\ infection\ rate = \frac{4\, s_0\, \rho\, u}{(1+\rho+u)^2}\left(1-\frac{1}{r}-\frac{\ln(r)}{r}\right)$$

Using again the approximations $u \cong \sqrt{4\rho r}$ and $\sqrt{\rho}+\frac{1}{\sqrt{\rho}} \cong 2$, we can write the following:

$$(16) \qquad Peak\ infection\ rate \cong \frac{2\, s_0 \sqrt{\rho}\sqrt{r}}{\left(1+\sqrt{r}\right)^2}\left(1-\frac{1}{r}-\frac{\ln(r)}{r}\right)$$

The peak infection rate is proportional to $s_0$. This means that if twice as many people are susceptible (immune-escaping variant), the peak will be twice as high. The height of the peak also depends on the latent period ($\rho$ is the ratio of the infectious period over the latent period). If the latent period is halved, then the peak infection rate will be multiplied by $\sqrt{2}$ = 1.4.

Further, the impact of a reduction in transmission is almost independent of $s_0$ and $\rho$ and decreases with $r$ as shown in Figure D2.

**Figure D2: impact of transmission reduction on the peak infection rate for different values of *r***

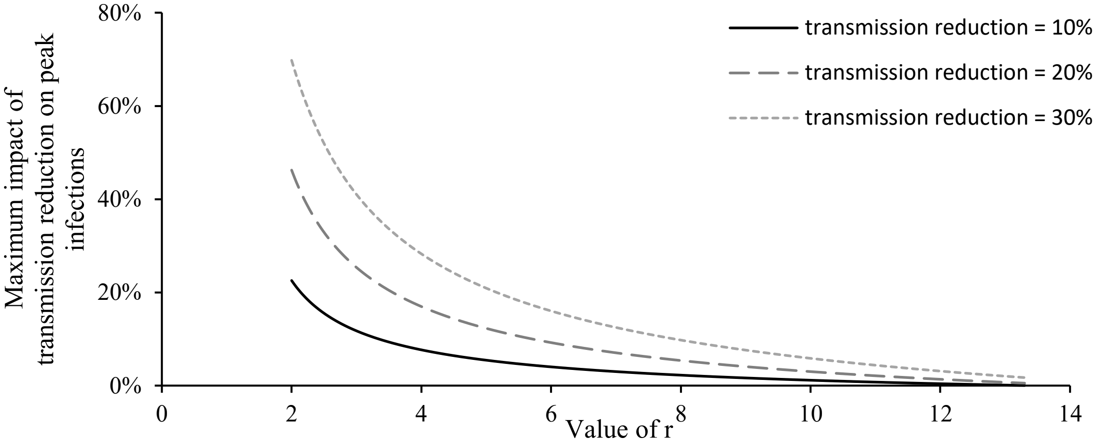
